# DNA methylation analysis of archival lymphoreticular tissues in Creutzfeldt-Jakob disease and the prevalence of vCJD infection in the UK

**DOI:** 10.1101/2022.06.16.22276495

**Authors:** Fernando Guntoro, Emmanuelle Viré, Chiara Giordani, Lee Darwent, Holger Hummerich, Jacqueline Linehan, Katy Sinka, Zane Jaunmuktane, Sebastian Brandner, John Collinge, Simon Mead

**Affiliations:** MRC Prion Unit at University College London (UCL), Institute of Prion Diseases, UCL, London W1W 7FF; STI & HIV Department and CJD Section, Public Health England National Infection Service, 61 Colindale Avenue, London NW9 5EQ; Division of Neuropathology and Department of Neurodegenerative Disease, UCL Queen square Institute of Neurology, Queen Square, London WC1N 3BG

## Abstract

The exposure of the UK and other European populations to bovine spongiform encephalopathy (BSE) prions caused human variant Creutzfeldt-Jakob Disease (vCJD) and a prolonged public health crisis. Throughout, a key question has been the prevalence of vCJD prion infection in the UK population. vCJD has several distinct features including immunohistochemically detectable abnormal prion protein (PrP) in peripheral lymphoreticular system tissues (LRS eg. tonsil, appendix). Surveys have detected abnormal PrP in the LRS of the UK population, but it remains unclear if these represent carriers of vCJD infection, some other form of prion infection, or another phenomenon altogether. Concern about the infectiousness of these possible carriers has been used to justify precautionary, expensive and ongoing health protection measures. Archival appendix samples are formalin fixed and paraffin embedded, a process that makes conventional assays of prion infection challenging. Here, we sought to use methylation array technology that assays >850,000 sites where chemically stable DNA modification occurs to develop a computational method to classify tissue samples by prion disease status. We assembled nearly 450 lymphoreticular tissue samples from patients with different prion diseases following biopsy or autopsy, and non-prion disease patients following tonsillectomy and appendicectomy, either frozen or processed as formalin fixed or formalin fixed paraffin embedded. DNA was extracted, bisulphite converted and assayed using Illumina Infinium Methylation EPIC (850K) BeadChips. Data were normalised and filtered, then analysed by case-control study, t-distributed stochastic neighbour embedding plots, and random forest classification methods. We show substantial differences in DNA methylation between prion diseases cases and controls, which can be exploited to classify LRS samples with reasonable levels of accuracy (82-97%). Archival appendix samples with abnormal PrP were most similar to, and classified with, control appendix samples, rather than prion disease samples; several interpretations are compatible with these findings.

## Introduction

Prions are proteinaceous infectious agents of humans and animals that propagate by the templated misfolding of normal cellular prion protein (PrP) into paired helical assemblies of misfolded forms of PrP(1). Prion diseases include sporadic Creutzfeldt-Jakob disease (sCJD) of humans, sheep scrapie, chronic wasting disease of cervids and the epidemic bovine spongiform encephalopathy (BSE) that devastated UK, and to a lesser extent other European cattle herds, in the early 1990s. Following the widespread dietary exposure of the UK and other populations to BSE prions and their human transmission as variant CJD (vCJD), a key question has been the prevalence of vCJD infection in the UK population(2). Clinically silent incubation times to acquired human prion disease can extend beyond 40 years and have powerful genetic determinants(3-5).

vCJD has several molecular and clinicopathological features that distinguish it from sCJD, notably, the presence of immunohistochemically detectable abnormal PrP in peripheral lymphoreticular tissues (LRS) like tonsil, spleen and appendix(6), findings that are distinct enough to be used for individual patient diagnosis(7). vCJD prions are also known to be present in human blood because the disease was transmitted by transfusion from donors who went on to die from vCJD(8, 9). These observations provoked concern about the infectiousness of blood (through transfusion) and LRS tissues, including gut associated lymphoid tissue (through general surgery) in people who might be subclinical carriers of vCJD. Whilst abnormal PrP and prions can be detected peripherally in sCJD, these findings are more variable and typically at lower levels(10).

Subclinical prion infection in the human population has been inferred by the detection of abnormal PrP associated with follicular dendritic cells by immunohistochemistry in LRS tissues taken from the healthy population and in vCJD patients prior to symptom onset(11-15). Prevalence surveys have been conducted using tonsillar or appendix tissue archived following surgical removal. The largest study of appendiceal tissues examined individuals within the birth cohort assumed to have been exposed to BSE (Appendix-II) and found abnormal PrP in 16/32441 samples(12). Definitive interpretation of this finding was hindered by the absence of a comparator group, unexposed to BSE, therefore Appendix-III examined archived appendiceal tissue obtained prior to 1980 when it was assumed that BSE exposure was insignificant and from those born after 1996, when it was assumed that BSE had been eliminated from the foodchain(13). Appendix III found 2/14692 appendix samples positive in the pre-1980 surgery cohort, both of these in a 1977-1979 window, and 5 within the 14,842 appendix samples from those born after 1996. The pre-1980 prevalence was nominally lower than that found in Appendix-II but was not statistically significantly different, and is seemingly out of keeping with an assumption of insignificant BSE exposure prior to 1980. One possible interpretation of these data is that there is an imperfect association between abnormal PrP in LRS tissue and vCJD prion infection, but other interpretations are also plausible(16).

A series of investigations were commissioned to help interpret the findings of abnormal PrP immunoreactivity in appendix tissue. The most obvious direct test of vCJD infection in appendix tissue is a bioassay by inoculation of mouse models, or use of amplification assays such as protein misfolding cyclic amplification (PMCA), but formalin fixation is known to reduce prion infection in tissues by 2 - 3 logs(17). Here, we sought to develop a new method of assessment based on genomic DNA methylation profiling to measure overall similarities between archival appendices with different PrP-reactivity status and those of known prion infection status. Gene expression is stably modified by DNA and/or chromatin structures including methylation of cytosine (CpG) sites. These covalent changes are dynamic, functional and disease specific and are known to be resistant to change during sample storage and tissue fixation(18). Highly multiplexed DNA methylation arrays have been developed that allow the discovery of sets of genomic sites that are altered by disease states, including prion diseases (19). Such approaches are currently revolutionising the classification of cancers, rendering possible more personalized treatments(20).

## Methods

### Samples, ethics, consent

Appendiceal and tonsillar tissues from patients with prion diseases and controls were archived at the MRC Prion Unit at UCL and obtained with informed consent for research studies. Frozen appendiceal tissue from prion disease patients was obtained at autopsy, whereas formalin fixed and paraffin embedded control appendiceal tissues were derived from those stored following the Appendix studies. Tonsillar tissue was obtained by biopsy or autopsy (in patients) or tonsillectomy (controls). Patients with sporadic and variant CJD were diagnosed using contemporary diagnostic criteria. London Queen Square Research Ethics Committee gave ethical approval for this work.

In some experiments part of the frozen tissue (FR) was formalin-fixed (FF) and part of it was formalin-fixed paraffin-embedded (FFPE) to test for the effects of sample processing on DNA methylation (FR, FF or FFPE referred to as tissue fixation ‘status’ hereafter). Formalin fixation was done according to standard procedures at the MRC Prion Unit at UCL by fixation in 10% buffered formalin for 72h. After formalin-fixation, samples were dehydrated in graded alcohols and then embedded in paraffin. Because of health and safety measures at the laboratories of the MRC Prion Unit at UCL, the FFPE processing was only done for control and not prion infected tissue.

### DNA extraction from appendix and tonsillar tissues

Possibly prion infected tissue was processed in a Biosafety Level 3 laboratory. Prior to DNA extraction, FR and FF samples were lysed using a Precellys Ribolyser and incubated in ATL lysis buffer (Qiagen) with proteinase-K overnight (50 μl proteinase K (from 20 mg/ml stock) Ambion) with mixing(19). For the FFPE tissues, dewaxing steps with Xylene were repeated twice and the tissue was digested overnight. DNA from these processed FR, FF and FFPE tissues was extracted with the Zymo Quick-DNA/RNA FFPE Kit according to the manufacturer’s instructions. DNA was cleaned using the Zymo Genomic DNA Clean & Concentrator. DNA concentration was determined with a Qubit Fluorometer. The quality of DNA extracted was also assessed using an Agilent TapeStation (as expected, the DNA integrity number (DIN) of FF and FFPE samples were lower than FR samples).

### Genome-wide methylation array processing

DNA restoration for the FFPE samples, bisulfite conversion, and methylation array processing were done by UCL Genomics at the Zayed Centre for Research into Rare Disease in Children. DNA from FFPE samples was restored using the Infinium HD FFPE DNA Restore Kit. Bisulfite conversion was performed using the Zymo EZ-96 DNA Methylation-Gold Kit. DNA samples from different tissue, disease status and processing groups were distributed evenly across batches and hybridized onto the Infinium MethylationEPIC BeadChips to measure the methylation levels of >850,000 CpG sites.

### Data preprocessing

Raw signal intensities were obtained from IDAT files using the minfi package in R. All samples were preprocessed using the preprocessIllumina() function, which performs background correction and control normalization which is comparable to Genome Studio implementation. Beta values were calculated from the transformed intensities using an offset of 100 (as recommended by Illumina). The beta values were then normalized using the BMIQ method implemented in ChAMP. To test for possible confounding batch effects within the dataset, the singular value decomposition (SVD) analysis implemented in ChAMP was used. Subsequently, a correction for significant batch effects was performed by fitting univariate, linear models using the removeBatchEffect() function in the limma package. The tonsil dataset was corrected for the type of material used (‘status’), whereas the appendix dataset was corrected for the EPIC array slide, based on the SVD analysis. Several filtering criteria were applied using champ.filter() function in the ChAMP package (Table 2) yielding 557,431 CpGs for further analyses.

**Table 1.** Characteristics of the samples used in the study. *Different fixation status samples are duplicates of those in the adjacent row. Where sample numbers in adjacent rows are unequal, the lower number is a subset of the larger number.

| Tissue | Diagnosis | Fixation status | n | Mean (SD) Age | %Female |
| --- | --- | --- | --- | --- | --- |
| Training Dataset |  |  |  |  |  |
| Appendix<br>(Total 155) | vCJD | Frozen | 10 | 31.5 (7.2) | 20.0% |
|  |  | Formalin-Fixed | 8* | 29.5 (6.6) | 25.0% |
|  |  | FFPE | - | - | - |
|  | sCJD | Frozen | 37 | 67.1 (9.5) | 51.3% |
|  |  | Formalin-Fixed | 38* | 67.1 (9.4) | 52.6% |
|  |  | FFPE | - | - | - |
|  | Control | Frozen | - | - | - |
|  |  | Formalin-Fixed | - | - | - |
|  |  | FFPE | 62 | 31.9 (12.6) | 41.9% |
| Tonsil<br>(Total 193) | vCJD | Frozen | 33 | 26.3 (8.0) | 42.4% |
|  |  | Formalin-Fixed | 32* | 25.9 (7.8) | 43.8% |
|  |  | FFPE | - | - | - |
|  | sCJD | Frozen | 7 | 46.7 (13.9) | 28.6% |
|  |  | Formalin-Fixed | 7* | 46.7 (13.9) | 28.6% |
|  |  | FFPE | - | - | - |
|  | Control | Frozen | 38 | 13.0 (12.6) | 39.5% |
|  |  | Formalin-Fixed | 38* | 13.0 (12.6) | 39.5% |
|  |  | FFPE | 38* | 13.0 (12.6) | 39.5% |
| Test Dataset |  |  |  |  |  |
| Appendix<br>(Total 96) | vCJD | Frozen | 8 | 32.8 (13.1) | 25.0% |
|  |  | Formalin-Fixed | 8* | 32.8 (13.1) | 25.0% |
|  | sCJD | Frozen | 12 | 65.0 (13.7) | 50.0% |
|  |  | Formalin-Fixed | 12* | 65.0 (13.7) | 50.0% |
|  | PrP-positive | FFPE | 20 | 30.1 (16.0) | 45.0% |
|  | PrP-negative<br>(Control) | FFPE | 36 | 33.7 (11.7) | 52.8% |

**Table 2.** Description of array processing and quality control of the data for further analyses. To filter low quality samples, a visual inspection of the beta distribution, an analysis of the fraction of CpG failed and sex matching were done. To filter low quality CpGs, the ChAMP package filter() function was used, which removes (1) probes with detection p-value over threshold of 0.01, (2) non-CpG probes, (3) SNP-related probes from Zhou et al. 2017(25), (4) multi-hit probes from Nordlund et al. 2013(26), and (5) probes located in chromosome X and Y.

| QC criterion | # of samples removed (*Training dataset*) | # of samples removed (*Test dataset*) |
| --- | --- | --- |
| Abnormal beta distribution / failed CpG fraction > 0.1 | 24 | 18 |
| Sex mismatch | 3 | 0 |
|  | # of CpGs removed (*Training dataset*) | # of CpGs removed (*Test dataset*) |
| Detection P-value above 0.01 | 167,042 | 173,553 |
| No CG start | 1612 | 1709 |
| Probes with SNPs | 80,891 | 81,576 |
| MultiHit | 14 | 18 |
| XY chromosome | 10,031 | 9965 |
| Remainder | 324 samples 606,501 CpGs | 78 samples 599,417 CpGs |
| Merged dataset | 402 samples 557,431 CpGs |  |

### Correlation analysis

Pairwise Pearson correlation was calculated for all appendix and tonsillar samples using all CpGs. Correlation statistics are summarised based on relevant comparisons between disease groups and sample status within appendix and tonsillar samples.

### Differential methylation analysis

The differential methylation analysis was performed using a linear regression model with the limma package. This was used to identify differentially methylated probes (DMPs) between vCJD and control FR tonsils. DMPs with a Bonferroni-adjusted P-values less than 0.05 were considered significant. A Manhattan plot was generated using an in-house script and a significance threshold was drawn at Bonferroni-adjusted threshold of 8.97 × 10^−8^. Quantile-quantile (QQ) plots were generated using an adapted version of the pQQ function of the haplin package and an area of 95% confidence level was shaded around the reference line. Kyoto Encyclopedia of Genes and Genomes (KEGG) pathways were analyzed using DAVID for genes with Adj. P Val. < 0.05 (21, 22).

### Unsupervised analysis

A t-distributed stochastic neighbour embedding (t-SNE) analysis was performed by selecting 10% of the most variably methylated CpGs (n = 55,743) using the Rtsne package. The following parameters were used: initial_dims = 86, theta = 0, max_iter = 2500, perplexity = 30. Perplexity values of 5 and 50 were also tested but showed no great discrepancy. The plots were then overlaid with relevant metadata: plate, sex, age and status.

### Machine learning classifier development

A published DNA methylation random forest classifier was adapted(20). The random forest algorithm is a supervised learning algorithm which relies on an ensemble of decision trees. To train the random forest classifier, the randomForest package was used. Feature selection to obtain the most important CpGs was done by applying the random forest algorithm to the beta values of pre-filtered 20,000 most variable CpGs (consider - all 557,431 CpGs). 10,000 trees were fitted and the selected features were ranked using the variable importance measure. The final random forest classifier was trained by fitting 10,000 trees using the beta values of the 20,000 CpGs selected during feature selection. The random forest model trained on the appendix Training dataset (see Table 1 for samples used) was used to predict the disease group classification in the Test dataset (Table 1) and produced a confusion matrix. The accuracy of the random forest model was evaluated by a threefold, nested cross-validation. The Training dataset was split into three equally sized parts, and in each iteration, two-thirds of the dataset were used to train a random forest classifier. The remaining one-third of the data were used for prediction using this random forest classifier.

## Results

We first analysed tonsillar tissue derived from vCJD and sCJD patients by tonsillar biopsy or at autopsy and from control tissue obtained at tonsillectomy. Demographic data are shown in Table 1 and quality control (QC) data in Table 2. Samples were necessarily imperfectly age-matched because of the distinct but overlapping age profiles of patients with prion diseases and/or those having tonsillectomy (Table 1). The purpose of this experiment was to test the hypothesis that a probable or definite prion disease diagnosis associates with a distinct profile of altered DNA methylation in lymphoreticular tissue, whilst correcting for the effects of age and sample processing. Samples that were divided and processed differently (FR, FF or FFPE) showed higher correlations than comparisons between samples from different individuals processed differently (Table 3). A genome-wide methylation association study demonstrated substantial differences (Figure 1a, b) and 184 statistically significant (Bonferroni corrected) differentially methylated sites in FR vCJD vs FR control tonsil (P_adj_<0.05, Figure 1, lambda=2.69) (Table 4). KEGG pathways analysed using DAVID identified two annotation clusters, both with relatively weak enrichment scores (Table 5) (21, 22). Significant terms were only found in Cluster 1 (B cell receptor signalling pathway, mTOR signalling pathway, VEGF signalling pathway, Sphingolipid signalling pathway; P <10^−3^).

**Table 3.** Correlation between paired frozen (Fr), formalin-fixed (FF) and FFPE samples, compared with unpaired samples and background correlation in each diagnosis group. Correlations between paired samples (ie those derived from the same original sample but processed differently) are stronger that the correlations between unpaired samples in all groups. However correlations between paired samples are not stronger compared to the background correlation levels between frozen samples. Background correlation levels of formalin-fixed samples is generally lower compared to background correlation levels of frozen and FFPE samples.

| Tissue | Diagnosis | Group | Mean r – Pairs (n) | Mean r – NonPairs (n) | Mean r – Fr (n) | Mean r – FF (n) | Mean r – FFPE (n) |
| --- | --- | --- | --- | --- | --- | --- | --- |
| Appendix | vCJD | Fr-FF | 0.9649<br>±0.0142 (6) | 0.9559 ±0.0140<br>(54) | 0.9759<br>±0.0129 (45) | 0.9535<br>±0.0166 (15) | NA |
|  | sCJD | Fr-FF | 0.9802<br>±0.0055 (37) | 0.9719 ±0.0149<br>(1369) | 0.9771<br>±0.0131 (666) | 0.9723<br>±0.0153 (703) | NA |
|  | Control | NA | NA | NA | NA | NA | 0.9734 ±0.0091<br>(1225) |
| Tonsil | vCJD | Fr-FF | 0.9817<br>±0.0105 (28) | 0.9758 ±0.0128<br>(900) | 0.9878<br>±0.0108 (496) | 0.9642<br>±0.0152 (406) | NA |
|  | sCJD | Fr-FF | 0.9784<br>±0.0234 (7) | 0.9598 ±0.0369<br>(42) | 0.9819<br>±0.0150 (21) | 0.9483<br>±0.0404 (21) | NA |
|  | Control | Fr-FF | 0.9774<br>±0.0159 (32) | 0.9721 ±0.0159<br>(1184) | 0.9914<br>±0.0032 (703) | 0.9534<br>±0.0217 (496) | 0.9832 ±0.0132<br>(703) |
|  |  | Fr-FFPE | 0.9922<br>±0.0090 (38) | 0.9868 ±0.0094<br>(1406) | NA | NA | NA |
|  |  | FF-FFPE | 0.9735<br>±0.0176 (32) | 0.9676 ±0.0181<br>(1184) | NA | NA | NA |

**Table 4.** Top 20 most differentially methylated positions (DMPs) from vCJD and Control in frozen tonsil. Adj.P.Value refers to a Bonferonni correction. Gene refers to nearest gene if within.

| Rank | CpG | Delta Beta | P.Value | Adj.P.Value | Chromosome | Gene |
| --- | --- | --- | --- | --- | --- | --- |
| 1 | cg16495530 | 0.096 | 5.54E-11 | 3.09E-05 | 11 | *UBASH3B* |
| 2 | cg17933764 | 0.145 | 6.54E-11 | 3.65E-05 | 17 | *TRAF4* |
| 3 | cg07433769 | 0.144 | 1.43E-10 | 7.99E-05 | 18 | *C18orf1* |
| 4 | cg04016841 | 0.100 | 4.23E-10 | 0.000236 | 11 | *DENND5A* |
| 5 | cg20105297 | -0.083 | 5.43E-10 | 0.000303 | 5 |  |
| 6 | cg13159505 | 0.106 | 9.87E-10 | 0.00055 | 17 | *RPTOR* |
| 7 | cg10478600 | 0.106 | 1.24E-09 | 0.000692 | 7 | *CARD11* |
| 8 | cg01598421 | 0.155 | 1.27E-09 | 0.000709 | 8 | *TG* |
| 9 | cg23873281 | 0.096 | 1.34E-09 | 0.000748 | 9 |  |
| 10 | cg12120178 | 0.102 | 1.35E-09 | 0.000755 | 3 | *LPP* |
| 11 | cg15894642 | -0.086 | 1.43E-09 | 0.000798 | 11 |  |
| 12 | cg10361659 | 0.149 | 1.50E-09 | 0.000835 | 13 | *RNASEH2B* |
| 13 | cg12626003 | -0.086 | 1.55E-09 | 0.000865 | 20 | *PHF20* |
| 14 | cg26264767 | 0.116 | 1.65E-09 | 0.000922 | 8 | *TG* |
| 15 | cg08437936 | 0.128 | 1.66E-09 | 0.000927 | 7 |  |
| 16 | cg19222059 | 0.109 | 1.81E-09 | 0.001009 | 8 |  |
| 17 | cg04018282 | 0.066 | 1.85E-09 | 0.00103 | 11 |  |
| 18 | cg11703639 | 0.123 | 1.92E-09 | 0.001069 | 14 | *LGMN* |
| 19 | cg25271915 | 0.109 | 2.03E-09 | 0.001134 | 9 | *SNX30* |
| 20 | cg21873971 | 0.102 | 2.13E-09 | 0.001187 | 3 | *PDIA5* |

**Table 5.** The results of KEGG pathway enrichment analysis for genes with differential methylation levels with Adj.P.Value<0.05. The enrichment score for each annotation cluster is based on the probability value used for each term member. “Count” represents the number of genes from the differentially methylated list found in each individual term (listed in column Nr of Genes in list). P. value is the result of a Fisher Exact test.

| **KEGG Pathway**<br>**Annotation cluster 1** | **Enrichment score 1.19** | **Count** | **Nr Genes in**<br>**list** | **P.Value** |
| --- | --- | --- | --- | --- |
| B cell receptor signalling pathway |  | 5 | 69 | 0.0013 |
| mTOR signalling pathway |  | 4 | 58 | 0.0075 |
| VEGF signalling pathway |  | 4 | 61 | 0.0086 |
| Sphingolipid signalling pathway |  | 5 | 120 | 0.0095 |
| Fc epsilon RI signalling pathway |  | 4 | 68 | 0.012 |
| Focal adhesion |  | 6 | 206 | 0.014 |
| Fc gamma R-mediated phagocytosis |  | 4 | 84 | 0.020 |
| Viral carcinogenesis |  | 5 | 205 | 0.054 |
| Pathway in cancer |  | 7 | 393 | 0.054 |
| Long-term depression |  | 3 | 60 | 0.065 |
| Glioma |  | 3 | 65 | 0.075 |
| Hepatitis B |  | 4 | 145 | 0.080 |
| PI3K-Akt signalling pathway |  | 6 | 345 | 0.091 |
| Chemokine signalling pathway |  | 4 | 186 | 0.14 |
| Choline metabolism in cancer |  | 3 | 101 | 0.16 |
| Proteoglycans in cancer |  | 4 | 200 | 0.16 |
| Rap1 signalling pathway |  | 4 | 210 | 0.18 |
| Leukocyte transendothelial migration |  | 3 | 115 | 0.19 |
| Natural Killer cell mediated toxicity |  | 3 | 122 | 0.21 |
| Wnt signalling pathway |  | 3 | 138 | 0.25 |
| MAPK signalling pathway |  | 4 | 253 | 0.26 |
| MicroRNA in cancer |  | 4 | 286 | 0.32 |
| Regulation of actin cytoskeleton |  | 3 | 210 | 0.43 |
| Ras signalling pathway |  | 3 | 226 | 0.47 |
| **KEGG Pathway**<br>**Annotation cluster 2** | **Enrichment score 0.66** | **Count** | **Nr Genes in**<br>**list** | **P.Value** |
| Toll-like receptor signalling pathway |  | 3 | 106 | 0.17 |
| Osteoclast differentiation |  | 3 | 131 | 0.23 |
| MAPK signalling pathway |  | 4 | 253 | 0.26 |

**Figure 1a.**
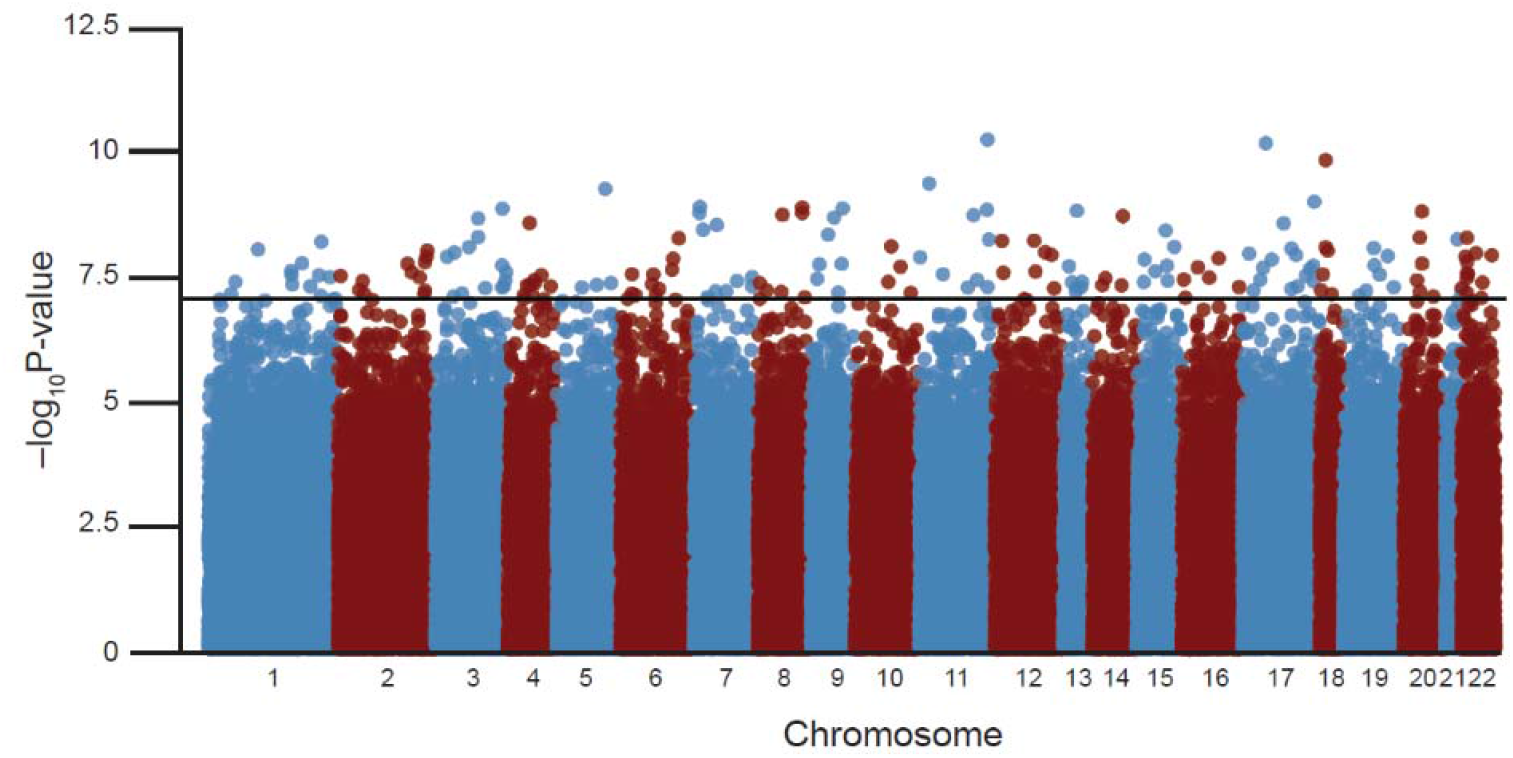
Manhattan plot vCJD tonsil vs case-control study.

**Figure 1b.**
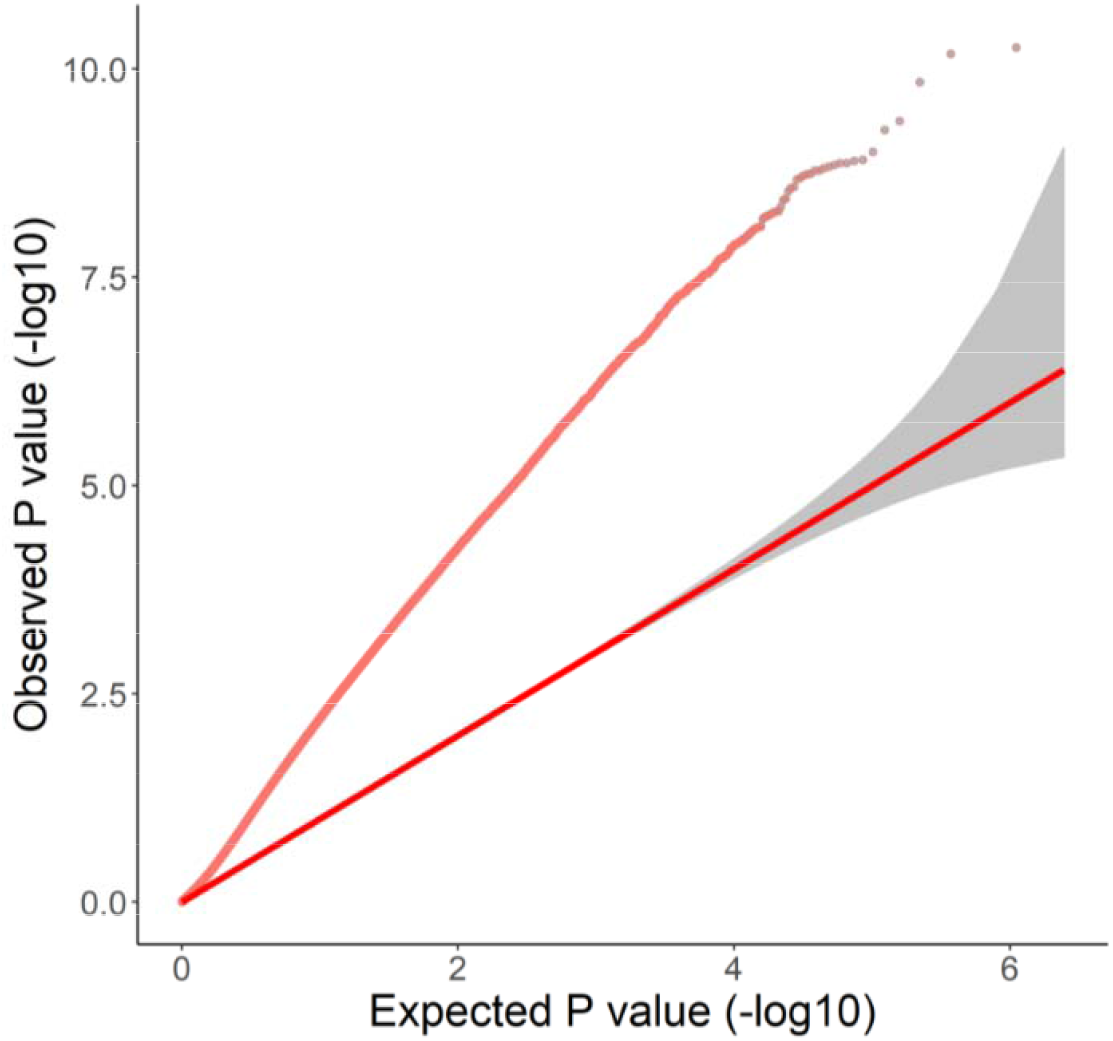
QQplot from the vCJD tonsil case-control study showing the relationship between observed P value and expected P value of 557,431 CpGs, showing an inflation lambda value of 2.69.

We used a t-SNE plot (Figure 1) to illustrate sample similarity by two-dimensional distance based on the most variable 10% of the DNA methylation probe dataset (Figure 2). These plots showed evidence of higher similarity between prion disease cases than between prion disease and controls, that was not confounded by age, sex or sample processing (FR, FF, or FFPE).

We classified tonsillar samples into vCJD, sCJD and control disease status using random forest machine learning (see Methods). 183 samples were analysed, 14 sCJD samples were classified correctly, as were 59/61 vCJD samples (errors: 1 as control, 1 as sCJD), and 89/108 controls (errors: 7 as sCJD and 12 as vCJD), with an overall accuracy of 0.885 (95% CI 0.830-0.928, P<2.2 × 10^−16^, one-sided test that accuracy was better than the prevalence of the most common category (in this case, control 108/183, 59%)). Overall misclassification error score based on 3-fold cross validation was 0.311 indicating a degree of overfitting in this classification.

We went onto analyse appendiceal tissues in a similar way (transparent symbols, Figure 3). As for tonsillar tissue, t-SNE showed a clear differences between prion disease samples and controls. In the training dataset 141 samples, 72/75 sCJD samples were classified correctly (errors: 2 as control, 1 as vCJD), 16 vCJD samples were classified correctly, and 48/50 control samples (errors: 2 as sCJD) with an overall accuracy of 0.965 (95% CI 0.919-0.988, P<2.2 × 10^−16^). Overall misclassification error score based on 3-fold cross validation was 0.092 suggesting a lower degree of overfitting compared with the tonsillar datasets.

**Figure 3.**
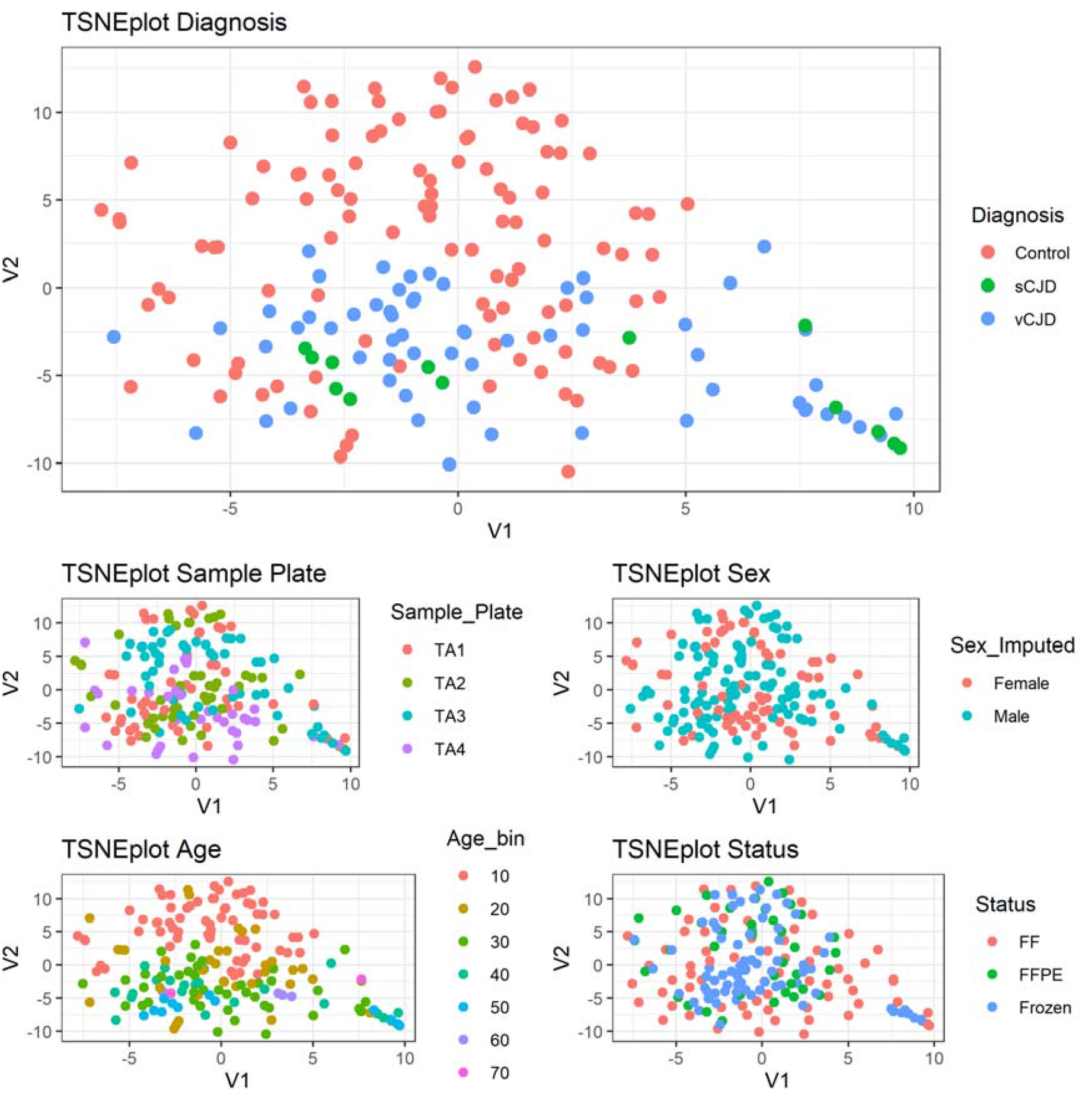
tSNE plots of DNA methylation profiles derived from tonsillar tissue samples. t-SNE plots illustrate the estimated DNA methylation profile similarities between pairs of samples on a 2D plot. More similar profiles are shown closer together on the plots. These five plots show the same sample locations with various overlay colours including diagnosis, sample plate, sex, age, and tissue processing status. Total sample size n = 183. Duplicate samples are shown connected in Supplementary Figure 2.

**Figure 3.**
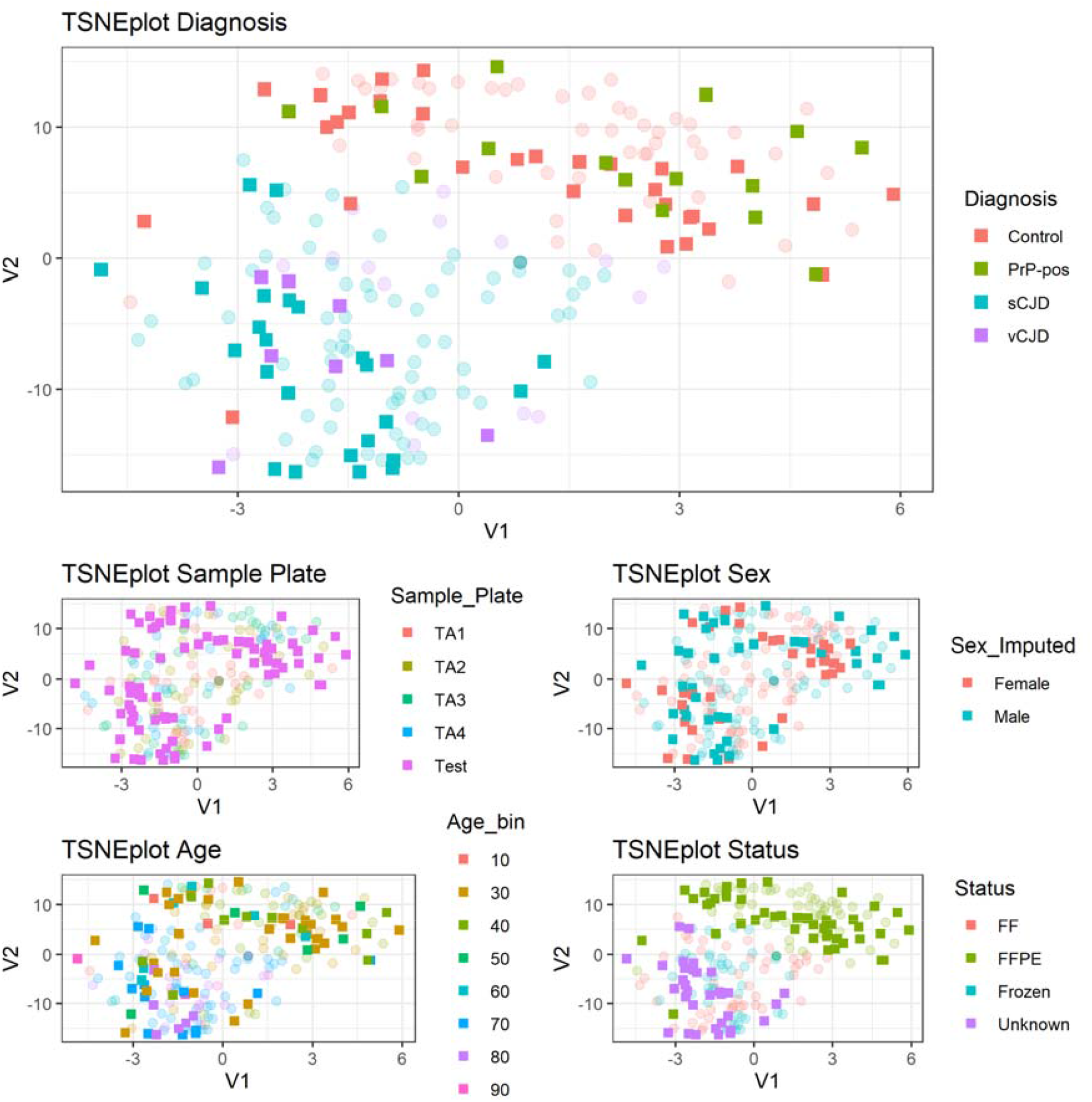
tSNE plot of DNA methylation profiles in including training (transparent) and test (opaque) appendix samples. Five plots show the same sample locations with various overlay colours including diagnosis, sample plate, sex, age, and tissue processing status. Test samples are shown in opaque colour on top of discovery/training samples in transparent colours. Total sample size n = 219. PrP-pos refers to Appendix II or III samples that showed abnormal PrP immunohistochemistry.

Encouraged by these findings we processed a blinded “test” plate containing 96 samples (Table 1, opaque symbols in Figure 3) comprising FR and FF pairs of known prion disease appendix samples, and FFPE samples from known controls (“PrP-negative”) and those of uncertain prion disease status with abnormal PrP immunoreactivity in follicular dendritic cells of the lymphoid follicle (“PrP-positive”). t-SNE showed that prion disease cases were most similar to the prion disease cases in the training set, whereas PrP-positive and PrP-negative samples were distributed evenly in a similar distribution to known control samples. In the test dataset 78 samples, 20/22 sCJD samples were classified correctly (2 as vCJD), all 10 vCJD samples were erroneously classified as sCJD, 30/31 PrP-negative samples were classified correctly (1 as vCJD), and 14/15 test PrP-positive samples were classified as controls. The overall accuracy was 0.821 (95% CI 0.717-0.898, P<1.2 × 10^−5^).

We went on to do sensitivity analyses that examined possible confounding effects. First, we considered the possibility that DNA methylation status in LRS tissue was determined by CJD disease severity in a general sense, rather than the presence of prions or abnormal PrP in the LRS tissue itself. In this circumstance we might expect that patients with more advanced disease would have more distinct DNA methylation profiles compared with controls than patients with less severe disease. We therefore labelled vCJD tonsil samples by whether they were obtained at post-mortem, or biopsy, and if at biopsy, then by the tercile of clinical duration (Supplementary Figure 1). We observed no obvious clustering of samples by disease severity.

In a second sensitivity analysis, we considered the possibility of confounding by tissue processing. We therefore did a genome-wide methylation association study comparing paired (n=38) tonsillar tissues processed as FF or FFPE, and separately comparing known vCJD appendix (n=8) to FFPE control appendix (n=36). We then repeated classification and t-SNE using a dataset that was filtered for the top ranked 50,000 DNA methylation probes in each genome-wide methylation association study. In both circumstances as above, 14/15 PrP-positive samples were classified as controls, and t-SNE showed PrP positive samples distributed amongst controls, overall accuracy remained similar (0.795-0.833), suggesting that tissue processing did not confound the classification to prion disease or control.

## Discussion

In this study we used DNA methylation analysis of lymphoreticular (LRS) tissues from patients with prion diseases and controls to develop a new diagnostic classification tool. We showed reasonable levels of accuracy of the method across different types of tissue and processing (82-97%). We went on to apply the classifier to tissues derived from the Appendix prevalence surveys, which came from people who were healthy from the point of view of prion disease, but showed abnormal PrP in follicular dendritic cells. These samples were distributed amongst controls in t-SNE plots and classified (14 control /1 vCJD) as control samples by random forest machine learning. These findings have several caveats and interpretations discussed below and assessed in sensitivity analyses.

Our interpretations must be constrained for several reasons. DNA methylation profiling and classification methods have proven to be extremely accurate in the context of ***large*** training sample collections derived from patients with known diagnoses, typically cancer. Cancer tissues show marked abnormalities of DNA methylation (eg. ∼3% CpG sites delta beta >0.15) because of the profound effects of neoplasia on the epigenetic status of the cell, whereas the case-control differences we observed in tonsil were remote from brain tissue that is most impacted by prion disease and therefore much more modest. The sample collections available for study were necessarily small due to the rarity of prion diseases, and the availability of diagnostic imaging and clinical criteria in life meaning that tonsillar tissues were sampled in only a small minority of cases of vCJD. The machine learning methods of classification and visualisations we used cannot be considered as robust, because typically thousands of samples are required before classification tools become stable(20).

We considered how effects of tissue fixation and processing might impact our results and used a DNA repair kit, despite the fact that there is good evidence for the stability of DNA methylation status in damaged DNA(23, 24). In sensitivity analyses we tested for the possibility of confounding by probes sensitive to processing methods by filtering out DNA methylation probes that we found were altered by tissue processing (when the same sample was divided and processed differently). Whilst we found no changes to the overall accuracy or classification of tissues of unknown status, we cannot exclude the possibility that the archival FFPE processing status of appendix tissue has a distinct DNA methylation profile that obscured the signature of prion infection.

A further important limitation is that we do not know the cause or timing of altered DNA methylation in peripheral lymphoreticular tissues of prion disease patients. This could relate directly to prion infection of the tissue, alternatively, DNA methylation changes might result from more non-specific factors related to the general ill-health of the patient, treatments given, poor nutritional state, hypoxia or infection around the time of death. Such non-specific effects or morbidity would not likely be present in the PrP-positive appendix samples and would therefore bias classification to control status. The Appendix study samples were most likely to have come from those with appendicitis (although obviously inflamed samples were excluded) (12), nevertheless other confounding diagnoses that led to appendicectomy might similarly have confounded the analysis. We have not been able to source lymphoreticular tissue samples from non-prion dementia patients that might be used to further explore this possibility. We addressed this in part by examining the effects of disease severity in vCJD on tonsillar DNA methylation, concluding this was not a strong effect, but these analyses do not eliminate the possibility of confounding by morbidities distant from the test tissue. In summary, our conclusions are caveated by the absence of a gold-standard training sample set of subclinical carriers of vCJD infection.

Three potential explanations of the Appendix prevalence data may be considered: (i) that some or all of the “positive” appendices are “false” positive in that the PrP immunohistochemistry finding is a consequence of a prion-unrelated phenomenon, (ii) that some or all of the positive appendix results are prion-related, but these are not specific to vCJD infection, perhaps indicating a phenomenon related to other types of prion disease, or (iii) that the assumptions about the start of the BSE epidemic were incorrect, and in fact, BSE exposure prior to 1980 and after 1995 was sufficient to result in the findings of Appendix-III. Whilst we found that the test appendix tissues were indistinguishable from control tissues, which might favour interpretation (i) above, it would be an over-interpretation to conclude that the Appendix studies showed a “false-positive” result, as other interpretations of our work are valid. Our work needs to be considered together with other programmes designed to help interpretation of the Appendix studies, and of course the accruing epidemiological evidence of the absence of clinical vCJD in the UK population (zero known cases in the last five years).

Single cell DNA methylation might have been insightful but was not possible on these samples due to the early stages of the technological advancements in single cell-methylomics and also with the confounding factor of the sample infectivity hampering single cell isolation. Future work might explore a role for DNA methylation in the molecular classification of CJD brain tissues. If an appendix sample collection from non-prion dementia patients might be acquired, analysis of this might assist in the interpretation of our current findings. With respect to other methods of classification of the Appendix tissues, bioassay is clearly problematic because of the effects of fixation, but other methods to directly amplify prion seeds by PMCA, observe prions by electron microscopy, or assay prion titre by novel methods could all be pursued.

## Data Availability

All data analysed in this report will be deposited here on publication

## Acknowledgements

This study was funded by the Department of Health’s Policy Research Programme and the Medical Research Council (UK). SM and JC are NIHR Senior Investigators. Some samples were acquired through funding by NIHR’s Biomedical Research Centre at University College London Hospitals NHS Foundation Trust. ZJ, and SB are supported by the Department of Health’s NIHR Biomedical Research Centre’s funding scheme to UCLH We are grateful to patients and their families for providing permission for research studies.

## Data Availability

All data analysed in this report will be deposited here on publication

---

**Supplementary Figure 1.**
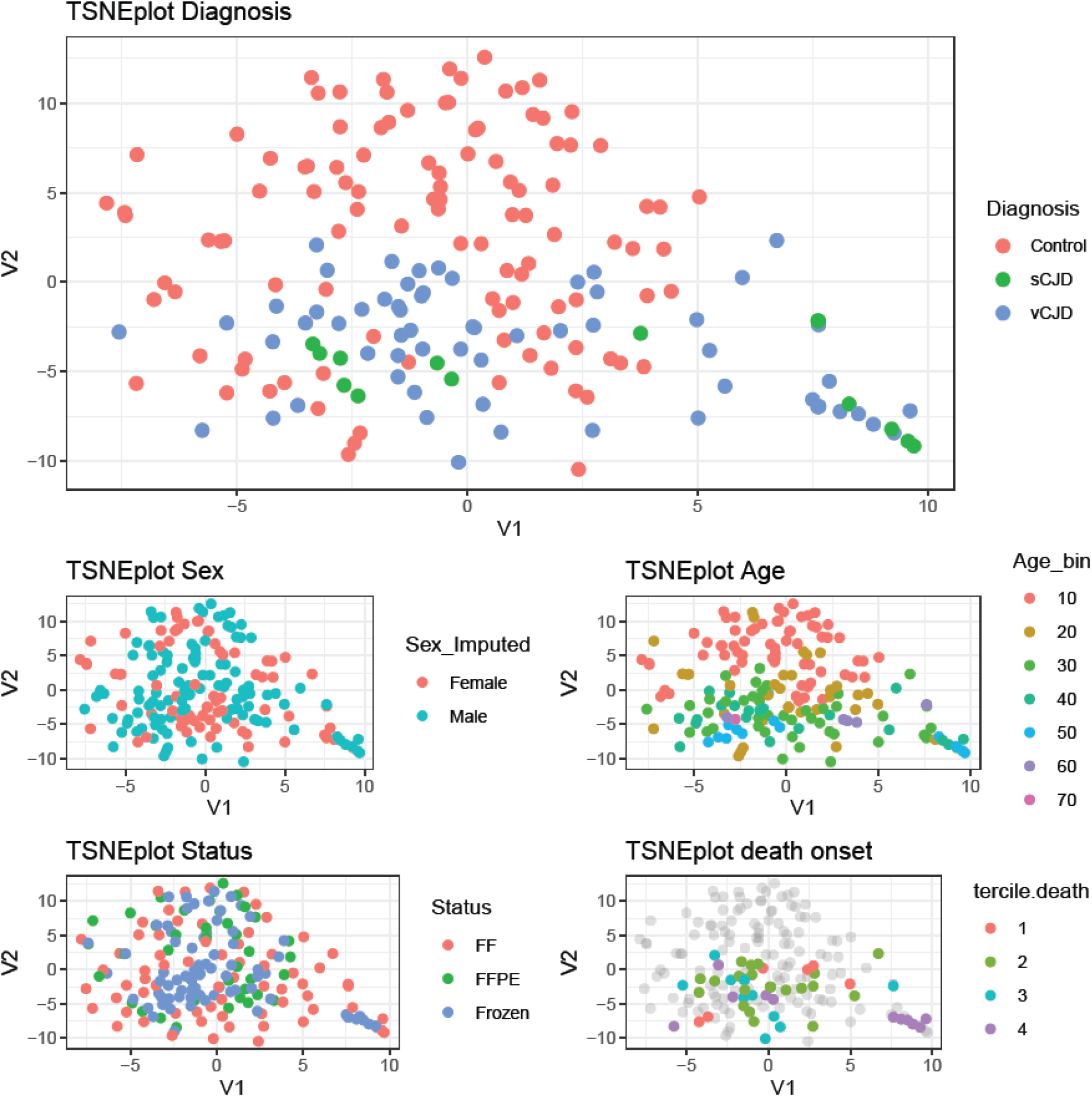
tSNE plots of DNA methylation profiles derived from tonsillar tissue samples. t-SNE plots illustrate the estimated DNA methylation profile similarities between pairs of samples on a 2D plot. More similar profiles are shown closer together on the plots. These five plots show the same sample locations with various overlay colours including diagnosis, sample plate, sex, age, and tissue processing status. Total sample size n = 183. For Terciles 1, 2, and 3, are the earliest, middle and late thirds of clinical duration. 4 = autopsy sample.

**Supplementary Figure 2.**
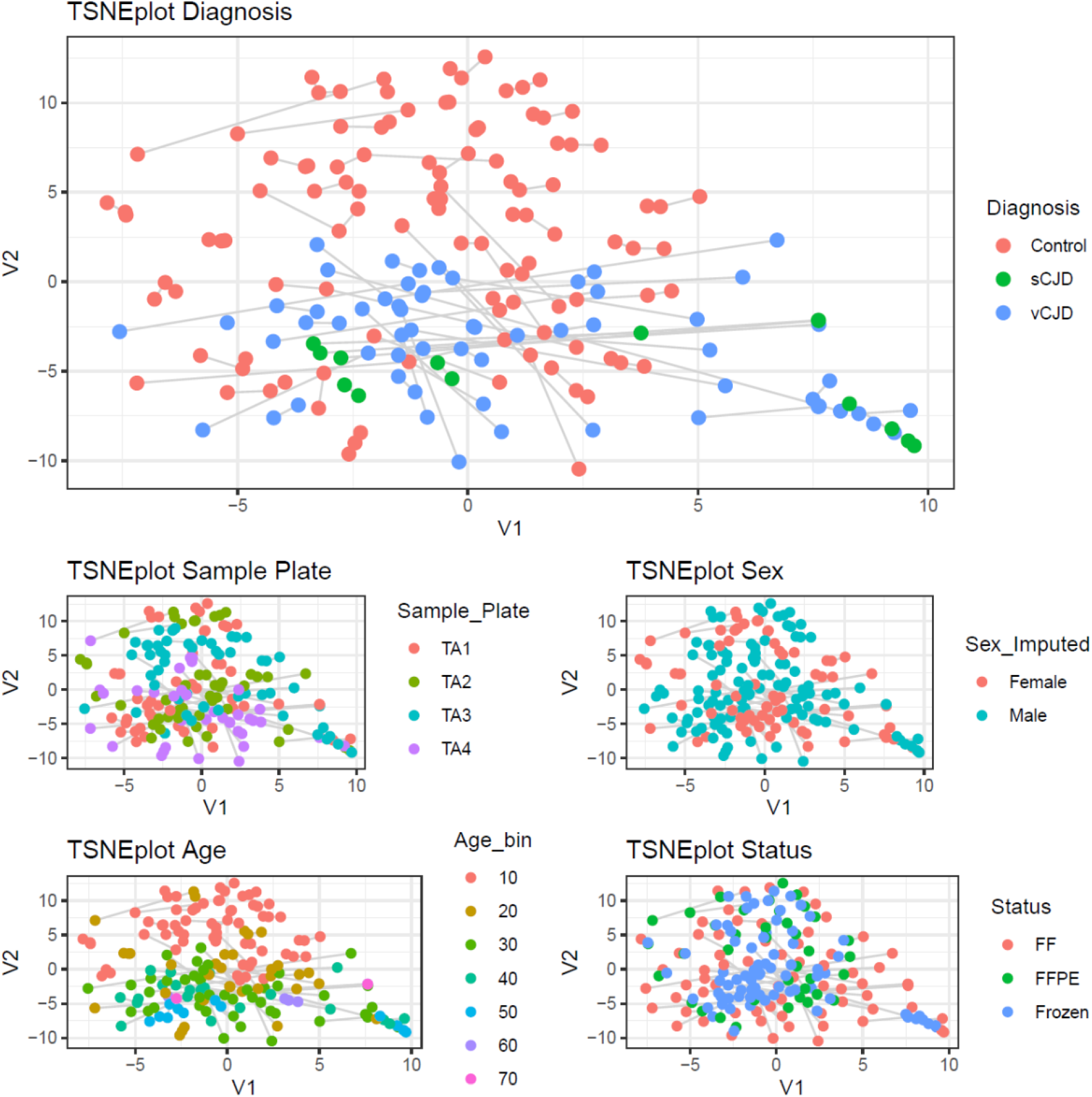
tSNE plots of DNA methylation profiles derived from tonsillar tissue samples. t-SNE plots illustrate the estimated DNA methylation profile similarities between pairs of samples on a 2D plot. More similar profiles are shown closer together on the plots. These five plots show the same sample locations with various overlay colours including diagnosis, sample plate, sex, age, and tissue processing status. Total sample size n = 183. Duplicate samples are shown connected.

